# Parameter estimation for the oral minimal model and parameter distinctions between obese and non-obese type 2 diabetes

**DOI:** 10.1101/2024.04.06.24305200

**Authors:** Manoja Rajalakshmi Aravindakshan, Devleena Ghosh, Chittaranjan Mandal, K V Venkatesh, Jit Sarkar, Partha Chakrabarti, Sujay K Maity

## Abstract

Oral Glucose Tolerance Test (OGTT) is the primary test used to diagnose type 2 diabetes mellitus (T2DM) in a clinical setting. Analysis of OGTT data using the Oral Minimal Model (OMM) along with the rate of appearance of ingested glucose (*R*_a_) is performed to study differences in model parameters for control and T2DM groups. The differentiation of parameters of the model gives insight into the behaviour and physiology of T2DM. The model is also studied to find parameter differences among obese and non-obese T2DM subjects and the sensitive parameters were co-related to the known physiological findings. Sensitivity analysis is performed to understand changes in parameter values with model output and to support the findings, appropriate statistical tests are done. This seems to be the first preliminary application of the OMM with obesity as a distinguishing factor in understanding T2DM from estimated parameters of insulin-glucose model and relating the statistical differences in parameters to diabetes pathophysiology.

**Highlights:**

- An established oral minimal model is used for estimation of parameters obese and non-obese type 2 diabetes.
- This is a novel attempt to use oral minimal model to co-relate the parameters for different diabetic groups using oral glucose tolerance test (OGTT) data.
- The data of different diabetes groups were utilised for model parameter determination and through statistical tests, distinctions in parameter distribution were identified.
- It was observed that obese diabetic group are more insulin resistant compared to non-obese diabetic group.

## 1 Introduction

Insulin glucose pathway (IG) is the biochemical regulatory pathway that helps to maintain a steady glucose level in the blood. Glucose is stored in liver cells in the form of glycogen. When the level of glucose falls in the blood (due to exercise or a long gap after the last meal), pancreatic α-cells secrete glucagon to release glucose (glycogenolysis) from the liver through the breakdown of glycogen until the glucose level rises to normal. When the level of glucose rises in the blood (after a meal), pancreatic β-cells secrete insulin to trigger the uptake of glucose by the peripheral tissue cells in the body, via the GLUT4 (Glucose transporter Type 4), until the glucose level falls back to normal. People suffering from diabetes mellitus (DM) need to regularly monitor their blood glucose level for hypogylcemia or hyperglycemia. Oral Glucose Tolerance Test (OGTT) is a commonly used test where fasting blood glucose is recorded and the subject is asked to ingest a certain amount of glucose dissolved in water and subsequently glucose and insulin readings are taken at varying intervals [1]. It is a highly sensitive test used for screening and diagnosis of pre-diabetes and type 2 diabetes mellitus (T2DM).

Machine learning (ML) models are used to identify and group data when adequate information about the data is not available. Features are extracted from the data in an abstract way in most of the ML models which makes it not easily explainable [2]. Clustering is an important ML technique employed to understand data through classification [3, 4, 5]. However, the factors leading to the classification may not be evident from the results. For example, data on diabetic patients may reveal distinct pathoclinical clusters [6], however, the causative factors may not be evident. Biological systems may also be modelled mathematically using ordinary differential equations (ODEs) utilising expert domain knowledge. Such models are typically parameterised, with the parameters playing specific roles in the model. The model parameters may be determined from available individual specific clinical data. For example, the insulin-glucose pathway may be mathematically modelled on such lines and the model parameters may be determined from available data on diabetic patients and also from a control group. The pool of parameter sets may then be studied to understand the causative factors with respect to the underlying model. Furthermore, the model may suggest specific internal processes which may be difficult to directly observe. This may be a significant advantage if the analytical results are verified to be reliable. In the current work, as features are not adequate and the available ones are time-series there is less scope of using abstract models and we focus on ODE models for insulin-glucose pathway.

ODE modelling started with the *minimal model* for modeling the intravenous glucose tolerance test (IVGTT). Bergman’s minimal model is a well accepted coupled model from the original model [7]. Another, simpler model with two coupled ODEs was introduced by Bolie [8]. A detailed feedback model with fourteen equations, introduced by Sturis et al. [9, 10], is regulated by three variables namely the amount of glucose in plasma and intercellular space, the amount of insulin in the plasma, and the amount of insulin in the intercellular space. The Hovorka model consists of a glucose subsystem to model absorption, distribution and disposal of glucose, an insulin subsystem (to model insulin absorption, distribution, disposal) and an insulin action subsystem (insulin action on glucose transport, disposal and endogenous production) [11].

Even though these models are widely used, a quantitative physiological model for insulin glucose pathway will be more suitable for epidemiological studies. This is achieved in the oral minimal model developed by Dallaman et al. [12] based on an oral glucose tolerance test (OGTT). A parameter *R*_a_, rate of appearance of oral glucose in plasma, is coupled with minimal model of glucose kinetics. Various *R*_a_ models have been developed based on gastric emptying data. Lehmann and Deutsch [13] proposed a trapezoidal gastric emptying function with a single compartment for the intestine whereas Elashoff et al. [14] considered exponential gastric emptying. Another linear model (Model 1 in [15]) with two compartments for stomach and a single compartment for intestine was proposed were the gastric emptying rate was constant. Another parametric description of *R*_a_ as piece-wise linear function was proposed [12] with known number of break times where there is shorter interval towards the beginning and longer intervals towards the end. A spline model and dynamic model were also proposed for an IVGTT model [12]. The Model 2 in [15], hereafter referred to as *R*_a_ model, is used as it is non-linear which is in accordance with the non-linear emptying of glucose (liquid phase).

In the Indian context, a great majority of people suffering from diabetes are non-obese, having low body mass index (BMI) [16]. This work is an investigation on the usability of the Oral Minimal Model (OMM), with the rate of appearance of ingested glucose (*R*_a_), for studying differences in obese and non-obese diabetes. The OMM is based on the Bergman minimal model [7] where glucose analysis is done using IVGTT. Identification of subject specific oral minimal models is carried out for the insulin-glucose pathway from the observed glucose and insulin readings in the OGTT data and inferring the rate of appearance of ingested glucose from the mathematical OMM. The model parameters of the participating subjects are studied towards suggesting differences in obese and non-obese diabetic physiology. Model identifiability is an important concern. Both the minimal model based on IVGTT and the rate of glucose appearance (Model 2 given in [15]) are globally identifiable when glucose and insulin are observed.

The problem with physiological models is the estimation of set of parameters both for an individual and a population. Some of the states in the model are not completely measurable and so estimation of parameters become an important step. The unknown states are characterised by approximating the states and outputs that are measurable. The following sections describe the mathematical description of the oral minimal model, theory, methodology followed by results and discussion.

## 2 Model under consideration

The oral minimal model with rate of appearance of ingested glucose, *R*_a_, is the computational model used in the work. This particular *R*_a_ model [15] was used as it is non-linear in accordance with the biphasic nature of gastric emptying of glucose. Initially the stomach contains the amount of ingested glucose. The gastric emptying rate then decreases to a minimum and rises back to *k*_*max*_ – this behaviour is also exhibited in the data used in this work. Also, the *R*_a_ model is modified by removing the solid phase glucose compartment from the stomach as it deals with a grinding rate parameter which is not relevant to our study as glucose solution is orally ingested. Equations 1, 2, 3 form the modified *R*_*a*_ model that is used in this work. The model equations are given as follows:

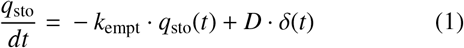

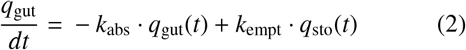

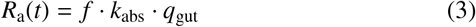

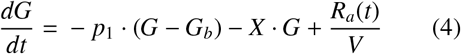

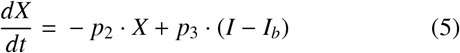

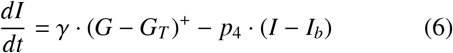

where *G, I* denotes blood glucose concentration and blood insulin concentration at time *t*, respectively. Here *X* is an auxiliary variable to model the time delay in insulin-dependent glucose uptake activity. The initial values of G and I are taken at the time when the glucose is orally ingested. Initial value of X is taken as 0 and (*G*− *G*_*T*_ )^+^ = (*G*− *G*_*T*_ ), if *G* > *G*_*T*_ and 0 otherwise. *G*_b_ is the baseline glycemia and *I*_b_ is baseline insulemia of the subject; *p*_1_ is the fractional glucose effectiveness i.e., the ability to promote glucose disposal and inhibit glucose production, *p*_2_ is the insulin-dependent increase in glucose uptake ability in tissue per unit of insulin concentration above *I*_*b*_, *p*_3_ is a scale factor for amplitude of insulin action and *p*_4_ is the first order decay rate for insulin in plasma, where *q*_sto_ is the amount of glucose in the stomach, δ(*t*) is the impulse function, *D* is the amount of ingested glucose, *q*_gut_ is the glucose mass in the intestine, *k*_empt_ is the rate of gastric emptying, *k*_abs_ is the rate constant of intestinal absorption, and *f* is the fraction of the intestinal absorption which actually appears in plasma.

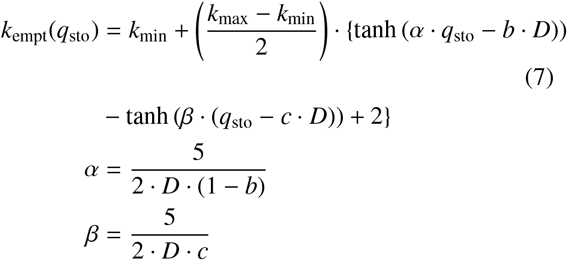

The gastric emptying rate, *k*_empt_ is a function of the amount of glucose in the stomach *q*_sto_ (Eqn 7). It equals *k*_max_ when the stomach contains the amount of the ingested glucose, *D*, then it decreases to a minimum value of *k*_min_. *b* is the percentage of the dose for which *k*_empt_ decreases at (*k*_max_-*k*_min_)/2. *c* is the percentage of the dose for which *k*_empt_ is back to (*k*_max_ − *k*_min_)/2.

## 3 Theory

### 3.1 Parameter sensitivity analysis methods

One of the crucial aspects of modeling is to understand how parameter values changes with model output. The parameter variations affect the output of a model when dealing with critical data specific to bio-physical pathways. The interpretation of parameter values in these pathways is important to understand the relationship to their physiological properties. To achieve this sensitivity analysis is performed. Two of the broad categories of sensitivity analysis are: local and global approach. To obtain local response of each parameter to a model local approaches are used which is specific to a region in parameter space. For global response of parameters a global approach which spans the entire parameter space is used.

Local sensitivity analysis is a simple technique which involves calculating the partial derivative of output with respect to the parameter considered. Since single point derivatives are taken this method is suitable for less complex cost functions. The parameter interactions are not considered. Global sensitivity analysis capture sensitivity over the entire parameter range. In biological systems most of the models are non-linear and variability and uncertainty of the inputs are to be considered over a wide range to estimate true model sensitivities. As global approach rely on sampling the parameter space this method becomes computationally expensive but the method captures the relative ranking of parameter influence.

Morris screening method is sampling based global sensitivity analysis technique. In this method elementary effects are averaged to rank the parameter importance by computing sensitivity measures. The individual effects of parameters are evident from the mean and variance of elementary effects. For detailed explanation of Morris screening algorithm refer Wentworth et al [17]. Fisher Information Matrix (FIM) is another measure to capture changes in parameter values which is based on calculated output sensitivities. FIM also gives an idea of how much information can be extracted by the parameters from the experimental data [18]. If FIM is non-singular then the parameters are identifiable [19]. Parameter sensitivity analysis is used to examine how sensitive a mathematical model responds to variations in its input variables. When every parameter in an ODE model is varied to understand model response, global sensitivity analysis is used. General formulation of a non-linear dynamic system is shown as:

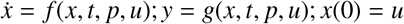

where 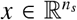 is the vector of *n*_*s*_ number of state variables, 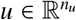 is the vector of *n*_*u*_ number of initial values of state variables, 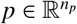 is the parameter vector of length *n*_*p*_ and 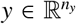 is the output vector of length *n*_*y*_. The first order sensitivity function is a sampling time dependent matrix *S* _*it*_(*t*_*k*_) such that the sensitivity value of *i*^*th*^ output variable concerning *j*^*th*^ parameter at time *t*_*k*_ is defined as:

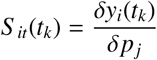

To determine which of the parameters have higher influence on the output, Morris screening method [17] is applied. It is a derivative-based approach to analyse sensitivity of a model function *h* : ℝ^*m*^→ ℝ to variations of *m* parameters *p*_*j*_ ∈ ℝ where *j* = 1, …, *m*. Under the assumption that *h* is at least once differentiable, partial derivatives 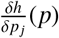 can be computed to define elementary effect directly. The elementary effect of the *j*^*th*^ parameter *p*_*j*_ on *h* is defined as:

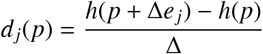

with step size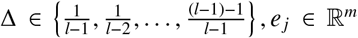, *e* _*j*_ ∈ ℝ^*m*^ is a small perturbation to parameter *p*_*j*_ and *l* can be any even number.

### 3.2 Parameter estimation methods

Parameter estimation aims to find unknown parameters in a computational model that may describe bio-physical pathways or phenomena. These unknown parameters are estimated using experimental data collected from well-defined and standard conditions. By minimizing the distance of theoretical function values and experimentally known data [20], the set of parameters in the model can be estimated. The parameters which are not directly measurable can be estimated using least squares or any other fitting methods to analyse the model quantitatively. This way, the behaviour of the model is captured effectively. Prediction models such as diabetes or pre-diabetes can also be built based on the analysis of estimated parameters.

A good parameter estimation algorithm is to be chosen when fitting a model to experimental data. The challenge is that no single optimal estimation technique exists for all models. Many different estimation methods have been developed so far to determine the best strategy for a given problem. The commonly used parameter estimation methods include maximum likelihood estimation [21] and Nelder Mead optimization [22]. Nelder Mead method is a numerical method in non-linear optimisation problems to find the minima of objective functions. Least square estimation is used in regression models, and maximum likelihood estimation is used in statistical models. Many other evolutionary methods of parameter estimation, namely genetic algorithms, also exists.

Our aim is to have an underlying well established model that is relevant and non-linear according to the context by estimating the set of parameters for each individual. This way the parameters obtained are clinically significant and each of the parameter has its associated clinical relevance.

## 4 Methodology

In this work the model parameters are estimated using the datasets described below. The sensitivity of the parameters are captured from the behaviour of a normal individual. The sensitivity indices and analysis methods used are also described in this section.

### 4.1 Datasets used for parameter estimation

Two sets of data are used in the study. Dataset-1 consists of OGTT data of 300 individuals of which 129 are normal subjects and others are T2DM. The outliers were removed and final data has 147 normal individuals and 116 diabetic individuals. These patients were asked to ingest 25g of glucose dissolved in 100mL of water after fasting for 8-12 hours. Sample data from dataset-1 are given in Table 1. FGLU indicates fasting glucose level. GLU ⟨*t*⟩ and INS ⟨*t*⟩ indicate glucose level and insulin level after time ⟨*t*⟩ of orally ingesting glucose beyond fasting glucose measurement (at *t* = 0), respectively. For example, GLU30 indicates glucose level after 30 min. The measurements are taken at times: 0, 30, 60, 90, and 120 minutes.

**Table 1:**
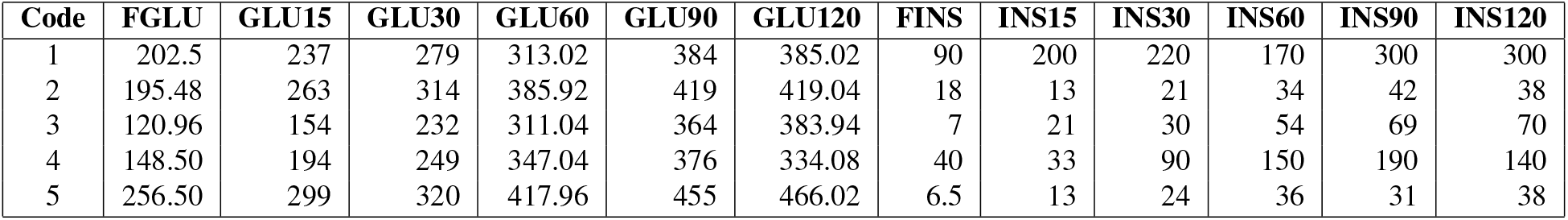
Sample data from Dataset 1

| Code | FGLU | GLU15 | GLU30 | GLU60 | GLU90 | GLU120 | FINS | INS15 | INS30 | INS60 | INS90 | INS120 |
| --- | --- | --- | --- | --- | --- | --- | --- | --- | --- | --- | --- | --- |
| 1 | 202.5 | 237 | 279 | 313.02 | 384 | 385.02 | 90 | 200 | 220 | 170 | 300 | 300 |
| 2 | 195.48 | 263 | 314 | 385.92 | 419 | 419.04 | 18 | 13 | 21 | 34 | 42 | 38 |
| 3 | 120.96 | 154 | 232 | 311.04 | 364 | 383.94 | 7 | 21 | 30 | 54 | 69 | 70 |
| 4 | 148.50 | 194 | 249 | 347.04 | 376 | 334.08 | 40 | 33 | 90 | 150 | 190 | 140 |
| 5 | 256.50 | 299 | 320 | 417.96 | 455 | 466.02 | 6.5 | 13 | 24 | 36 | 31 | 38 |

Dataset-2 consists of 40 T2DM patient data collected from a community clinic with 21 non-obese subjects (BMI ≤ 25) and 19 obese subjects (BMI>25) who underwent OGTT (age range 30-55). The outliers were removed and final data has 15 non-obese diabetic individuals and 14 obese diabetic individuals. These patients were asked to ingest 75g of glucose dissolved in 100mL of water after fasting for 8-12 hours. The sample data from dataset-2 are shown in Table 2. The dataset-2 consists of the following columns: Sex, Age, BMI, waist circumference (WC), Weight, Height and measurements of insulin and glucose at various time points. The column names are similar to Dataset-1 with only change in time points. The measurements in Dataset-2 are taken at times: 0, 15, 30, 45, 60, 90, and 120 minutes. For example, INS45 indicates insulin level after 45 min. All participants gave informed consent and the study was approved by Institutional Human Ethics Committtee of CSIR-IICB.

**Table 2:** Sample data from Dataset 2

| Code | SEX | BMI | WC | Weight | Height | FINS | INS15 | INS120 | FGLU | GLU15 | GLU120 |
| --- | --- | --- | --- | --- | --- | --- | --- | --- | --- | --- | --- |
| DM-24 | F | 23.8 | 82 | 47.4 | 141 | 0.64 | 0.47 | 6.38 | 167.80 | 192.50 | 400.69 |
| DM-09 | F | 21.7 | 80 | 47.1 | 146 | 5.30 | 10.00 | 25.17 | 159.73 | 189.39 | 398.13 |
| DM-28 | F | 24.2 | 86 | 51.5 | 146 | 2.42 | 3.42 | 31.06 | 145.86 | 146.23 | 245.98 |
| DM-34 | F | 24.6 | 84 | 50.4 | 143 | 7.59 | 8.08 | 36.48 | 140.83 | 179.63 | 344.74 |
| DM-06 | F | 20.8 | 75 | 45.9 | 147 | 7.24 | 10.43 | 36.63 | 154.75 | 178.68 | 309.04 |

### 4.2 Parameter Sensitivity

The parameter importance index δ_*l*_ [23] can be calculated using the sensitivity of the *l*-th parameter *p*_*l*_ to the output vector as follows:

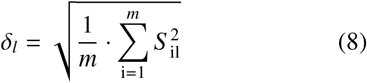

where, *m* is the number of parameters of the model. Collinearity index is calculated to analyse whether pairs of parameters have same influence on output vector. If two parameters have same effect on output vector they are likely to be not identifiable. For this normalised sensitivity matrix is calculated as follows:

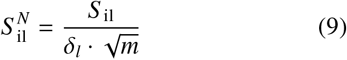

The submatrices are calculated as *m*× *l* with first *l* columns of *S* _*N*_ with respect to sensitivities of first *l* parameters. Then the smallest eigenvalue λ_*l*_ of each *l* × *l* matrix is computed and the index is defined as follows:

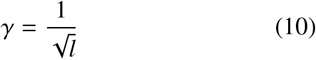

The parameters *p*_1_ to *p*_*l*_ are linearly dependent if the collinearity index is larger.

### 4.3 Parameter estimation

The parameters of the model in section 2 need to be estimated using the datasets described in section 4.1. This is described as parameter optimization problem that minimise the error between outputs obtained from solving ODE and the observed data. The methods are implemented and coded in python framework using the Scipy packages odeint(), minimize() and dist() [24]. Parameter estimation runs were performed for each individual data separately. Different estimation techniques were tried out and Nelder-Mead [22] gave a better fit of observed data. A weighted error function is defined which works with Nelder-Mead optimization. In the OMM estimation, the initial time intervals are crucial and so higher weights are given to initial time intervals (15, 30, 60 mins) compared to the later ones.

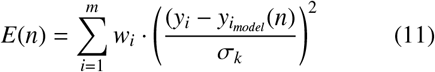

The weighted error function E(n) is referred in Eqn. 11, where *y*_*i*_ are data points, 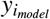 are simulated points and *w*_*i*_ are vector of weights associated with each timepoint.

## 5 Results

### 5.1 Sensitivity indicators

The implication of doing sensitivity analysis were to understand the varying levels of sensitivity among the parameters. The parameters which were less sensitive were replaced with nominal values but the overall fitting were not satisfactory. So it is understood that the small variation in less sensitive parameters is not of importance. But these parameters cannot be ignored altogether as they may be significant for larger variation among the disease sub categories. The range of these parameters may change over sub-groups. For less sensitive parameters smaller variation is not considered as they vary significantly across subgroups.

Morris screening algorithm were used to analyse the sensitivity for each individual data. The parameters which are highly sensitive are *k*_min_, *k*_max_, *k*_abs_, *f, p*_1_ and *G*_*b*_. The sensitivity order of parameters for all the groups remained mostly similar. The parameter importance index (Eqn. 8) and collinearity index (Eqn. 9, 10) were computed for all the patients in each dataset and the order is similar for most of the individuals. The first six parameters in sensitivity importance order are similar across all individuals and the later ones are slightly different in order across obese and non-obese T2D individuals, but largely the same. The ordering of parameters based on parameter sensitivity index are shown in Figure 1a. *k*_min_ is the parameter to which the output vector is most sensitive. The next set of important parameters are *k*_max_, *k*_abs_, *f, p*_1_ and *G*_*b*_. The least important parameters are *I*_*b*_ and *p*_2_. The collinearity index graph (Figure 1b) indicate that the first eight parameters in order are not strongly collinear with each other. This observation is mostly similar across all individuals with variations only among parameters which are less sensitive. The parameters which are collinear are found to be less sensitive to the output.

**Figure 1.**
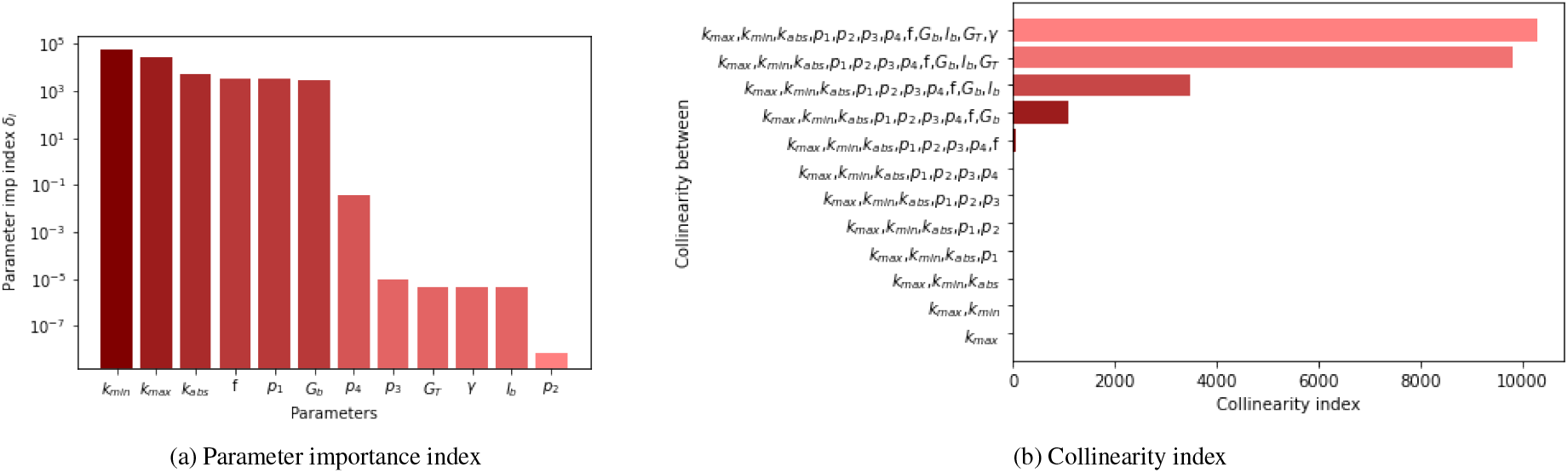
Parameter order in accordance with parameter importance index and collinearity index

### 5.2 Results of parameter estimation

The estimation considered 12 different parameters. The values of initial parameters are mainly from the literature and are used as required to match the unit of measurements. The initial values of insulin and glucose are taken at time point 0 (the time at which glucose is ingested) in the data and are not estimated. The model is estimated for parameter values for each of the groups separately. Simulation results of two random data samples from each of control and diabetic groups are shown in Figure 2. Sample results of two random individual’s data from obese and non-obese diabetic groups are shown in Figure 3.

**Figure 2.**
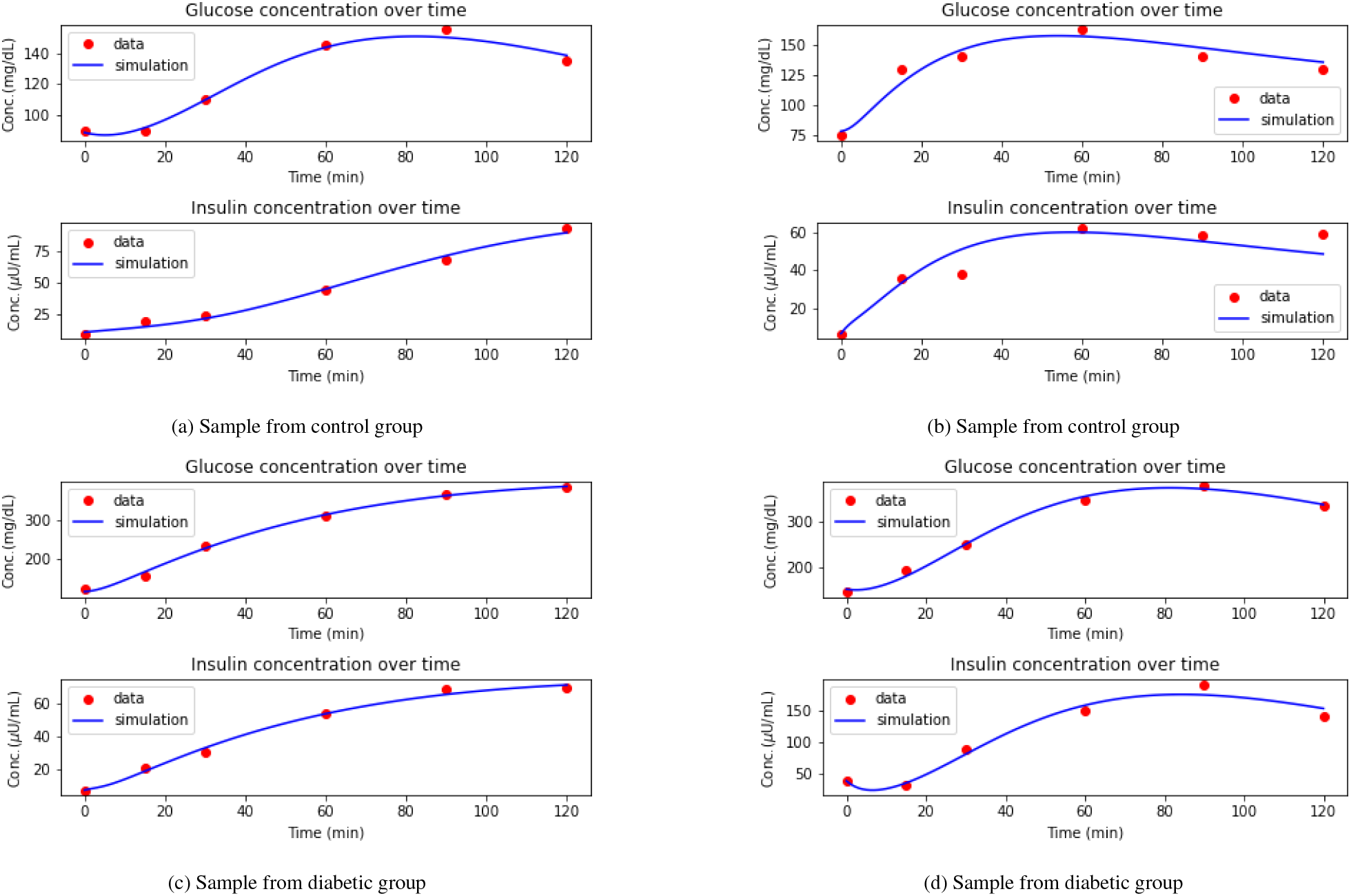
Sample simulation results for control and diabetic group

**Figure 3.**
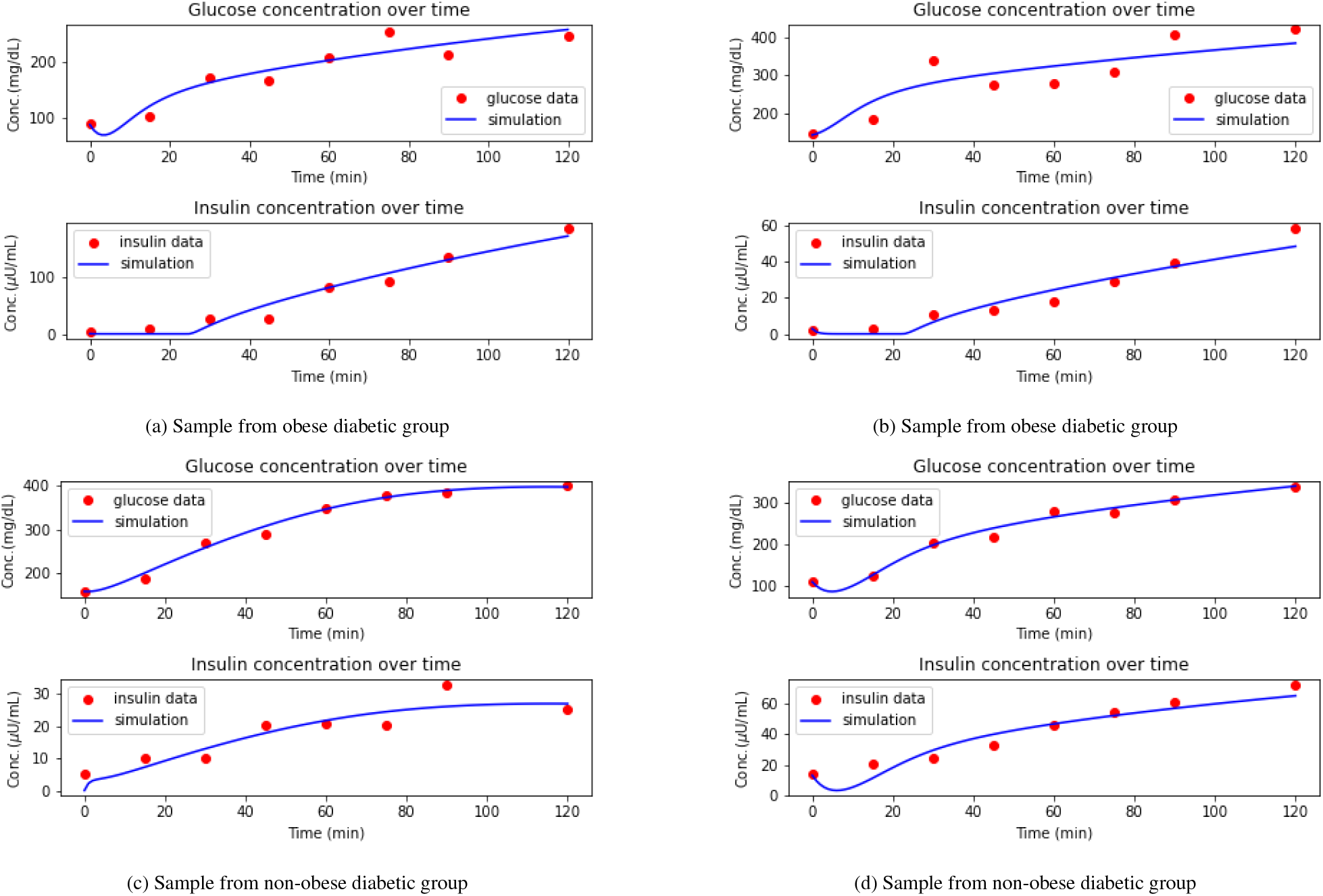
Sample simulation results for obese and non-obese diabetic group

### 5.3 Distribution of parameters and statistical test

The median values of parameters for control and diabetic groups are shown in Figure 4 and corresponding box plots are given in Table 3. For obese diabetic and non-obese diabetic group the box plots are shown in Figure 5 and median values are shown in Table 4. The box plots of parameters which are distinctive are only shown. Each parameter has different distribution and the statistical test are chosen based on the corresponding distribution. For instance in control and diabetic group *k*_*min*_ follows Weibull distribution (Table. 3) and a common non-normal distribution test, known as Mann Whitney U test is used. Similarly based on parameter distribution appropriate statistical test are chosen and pvalue is determined with significance level α = 0.05. The distribution followed and tests used for control and diabetic group using Dataset-1 are shown in Table 3 and for obese and non-obese diabetic group using Dataset-2 are shown in Table 4.

**Table 3:** Comparison of parameters in control and diabetic group

| Parameter | Control group | Diabetic group | p-value | Distribution | Statistical test | Distinct feature |
| --- | --- | --- | --- | --- | --- | --- |
| $k_{\max}$ [1/min] | 0.0179 | 0.0269 | 0.000143 | lognormal | Independent ttest | Yes |
| $k_{\min}$ [1/min] | 0.00137 | 0.00751 | 7.49e-27 | Weibull | Mann Whitney U test | Yes |
| $k_{\text{abs}}$ [1/min] | 0.0289 | 0.0208 | 0.0901 | lognormal | Independent ttest | No |
| $p_1$ [1/min] | 0.0609 | 0.0208 | 2.45e-26 | Weibull | Mann Whitney U test | Yes |
| $p_2$ [1/min] | 0.0016 | 0.0016 | 0.579 | Weibull | Mann Whitney U test | No |
| $p_3$ [(L/$\mu\text{mol}$)min<sup>-2</sup>] | 1.5e-07 | 1.5e-07 | 0.618 | Weibull | Mann Whitney U test | No |
| $p_4$ [1/min] | 0.154 | 0.863 | 5.61e-26 | Weibull | Mann Whitney U test | Yes |
| f [dimensionless] | 0.756 | 0.891 | 0.000125 | beta | Mann Whitney U test | Yes |
| $G_b$ [$\mu\text{mol/L}$] | 4.76e+03 | 7.3e+03 | 1.06e-12 | beta | Mann Whitney U test | Yes |
| $I_b$ [$\mu\text{mol/L}$] | 2.22e-11 | 2.22e-11 | 0.232 | Weibull | Mann Whitney U test | No |
| $G_T$ [$\mu\text{mol/L}$] | 4.24e+03 | 3.57e+03 | 0.364 | gamma ($\Gamma$) | Mann Whitney U test | No |
| $\gamma$ [min<sup>-2</sup>] | 2.8e-08 | 1.27e-08 | 2.15e-09 | Weibull | Mann Whitney U test | Yes |

**Table 4:** Comparison of obese diabetic and non-obese diabetic group

| Parameter | Obese diabetic (median) | Nonobese diabetic (median) | p-value | Distribution | Statistical test | Distinct feature |
| --- | --- | --- | --- | --- | --- | --- |
| $k_{\max}$ [1/min] | 0.0166 | 0.0182 | 0.561 | gamma ($\Gamma$) | Mann Whitney U test | No |
| $k_{\min}$ [1/min] | 0.00563 | 0.00252 | 0.000925 | t | Mann Whitney U test | Yes |
| $k_{\text{abs}}$ [1/min] | 0.00722 | 0.00141 | 1.16e-06 | exponential | Mann Whitney U test | Yes |
| $p_1$ [1/min] | 0.0565 | 0.15 | 0.000141 | Weibull | Mann Whitney U test | Yes |
| $p_2$ [1/min] | 0.00153 | 0.00808 | 0.451 | Weibull | Mann Whitney U test | No |
| $p_3$ [(L/$\mu\text{mol}$)min<sup>-2</sup>] | 2.48e-07 | 5.97e-08 | 0.186 | Weibull | Mann Whitney U test | No |
| $p_4$ [1/min] | 0.91 | 0.921 | 0.146 | Weibull | Mann Whitney U test | No |
| f [dimensionless] | 0.826 | 0.784 | 0.0259 | Weibull | Mann Whitney U test | Yes |
| $G_b$ [$\mu\text{mol/L}$] | 7.31e+03 | 5.04e+03 | 0.0292 | beta | Mann Whitney U test | Yes |
| $I_b$ [$\mu\text{mol/L}$] | 5.34e-12 | 5.01e-13 | 0.533 | beta | Mann Whitney U test | No |
| $G_T$ [$\mu\text{mol/L}$] | 4.58e+03 | 9.27e+03 | 0.102 | gamma ($\Gamma$) | Mann Whitney U test | No |
| $\gamma$ [min<sup>-2</sup>] | 1.65e-08 | 3.54e-09 | 0.134 | gamma ($\Gamma$) | Mann Whitney U test | No |

**Figure 4.**
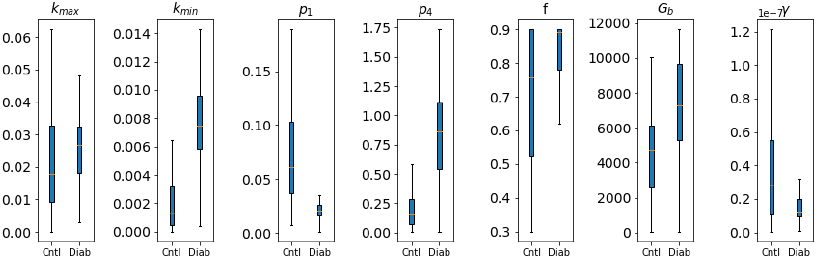
Box plots of parameters for control and diabetic subjects

**Figure 5.**
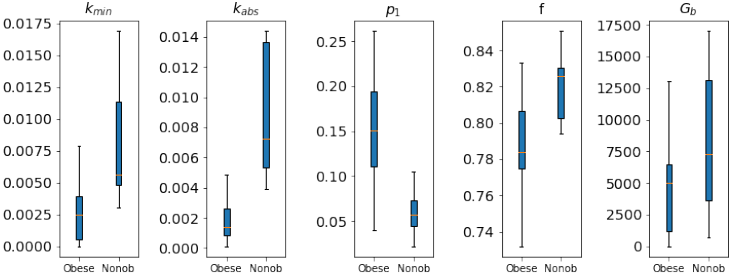
Box plots of parameters for obese and non-obese diabetic

## 6 Discussion

Our results emphasis the difference in parameter values in different T2DM groups. From Table 3 the control group differs from T2D groups based on parameters *k*_*max*_, *k*_*min*_, *f, p*_1_, *G*_*b*_, *p*_4_, and γ. The model parameters are conforming with the previously published results for diabetic patients which are described in detail in this section. Maximum and minimum amount of glucose emptying (*k*_*max*_ and *k*_*min*_) for diabetic group is higher. In support of this observation it can be argued that rapid gastric emptying is a frequent and important for diabetic complication [25]. To improve the postprandial glycemic control in these patients a slowing gastric emptying rationale may be considered. The fraction of intestinal absorption (f) which actually appears in plasma is higher for diabetic group. In patients with T2DM an increase of the Sodium/GLucose coTransporter 1 (SGLT1) protein and its mRNA in the enterocytes of small intestine were found which is involved in increased glucose absorption through the apical membrane [26]. The insulin independent rate of glucose uptake (*p*_1_) which also represent glucose effectiveness, is more in control group than in diabetic group. This may be attributed to insulin resistance among diabetes group. Basal level of glucose (*G*_*b*_) is higher in diabetes group which indicate pre-diabetes or diabetes which is expected. As this parameter is less sensitive, the variation among subgroups are larger but less significant. Smaller changes in basal level of glucose do not change the model behaviour. The decay rate for insulin (*p*_4_) is significantly higher in diabetic group. The excess insulin produced is rapidly attenuated due to acute induction of mitochondrial super-oxide production [27]. The rate of β-cells release of insulin (γ) after the oral glucose intake is lower in diabetic group.This may be due to decrease in insulin sensitivity and secretion caused by delay in glucose peak time [28].

The analysis of data using the model for obese and non-obese indicated that the parametric range is different for the two groups. From Table 4, it can be observed that the obese diabetic group differs from non-obese diabetic groups based on parameters *k*_*min*_, *k*_*abs*_, *f, p*_1_ and *G*_*b*_. The five estimated parameters are significant for obese and non-obese T2D groups. The minimum level of gastric emptying (*k*_*min*_) is higher for obese i.e., from that point the emptying rate increases to *k*_max_, not reducing any further. Obese subjects have more rapid emptying rate for solids than non-obese subjects [29]. Absorption rate in the intestine (*k*_*abs*_) is higher for obese diabetic. The body surface area for obese subjects are larger and therefore the absorption rate increases. The insulin independent rate of glucose uptake (*p*_1_) is higher for non-obese diabetic. This observation is associated with adiposity with declining glucose tolerance. During increased insulin resistance this mechanism help preserve glucose uptake [30]. The fraction of intestinal absorption ( *f* ) is higher for obese group. This has the same explanation for why *k*_abs_ is higher for obese diabetic group. Basal level of glucose (*G*_*b*_) is higher for obese diabetic group. These significant changes in parameter values for obese and non-obese indicates higher risk of diabetes as it causes both insulin resistance and β-cell dysfunction.

The differences in obese and non-obese diabetes group has to be discussed further as it is being considered as a novel category of diabetes. The degree of change and the direction of parameter values to increase or decrease in diabetes risk is studied. The risk of T2D increases for obese group with an increase in the value of parameter *k*_*min*_ as compared to non-obese group. The insulin independent rate of glucose uptake (*p*_1_) is 2.6 times lower in obese diabetic group indicating higher risk in obese diabetes. The absorption rate in the intestine (*k*_*abs*_) in obese group is 5 times higher compared to non-obese group indicating higher levels of glucose in non-obese diabetes group. This is also supported by higher fraction of intestinal absorption (f). The higher basal level of glucose (*G*_*b*_) in obese diabetic group indicate more insulin resistance. From the comparison it can also be concluded that obese group is more insulin resistant than non-obese group.

## 7 Conclusion

In this study we adapted a published model to determine the significant changes in parameter values for healthy and diabetic groups and further among obese and non-obese diabetes group. This is a first attempt to use oral minimal model to co-relate the parameters for different diabetic groups using OGTT data. The parameter values which were sensitive among groups were co-related to the known biological findings. The obese diabetic group are more insulin resistant whereas nonobese diabetic group are less insulin resistant. As nonobese T2DM is becoming more recognised a model to identify the differences in insulin secretion and glucose absorption among these phenotype are important. Using this model estimation more relevant studies can be conducted which pave way to precision therapy in T2DM.

## Data Availability

All data produced in the present study are available upon reasonable request to the authors

## Acknowledgement

The authors gratefully thank the associates of K V Venkatesh for collecting the insulin-glucose time series data. This work was partly funded by MoE, Government of India.

## References

[1] U. D. of Health, H. Services, Diabetes & prediabetes tests, https://www.niddk.nih.gov/health-information/professionals/clinical-tools-patient-management/diabetes/diabetes-prediabetes?dkrd=hisce0124 (2020).

[2] N. Burkart, M. F. Huber, A survey on the explainability of supervised machine learning, J. Artif. Intell. Res. 70 (2021) 245–317.

[3] M. R. Karim, O. Beyan, A. Zappa, I. G. Costa, D. Rebholz-Schuhmann, M. Cochez, S. Decker, Deep learning-based clustering approaches for bioinformatics, Briefings in Bioinformatics 22 (1) (2020) 393–415. doi:10.1093/bib/bbz170.

[4] W. T. Li, J. Ma, N. Shende, G. Castaneda, J. Chakladar, J. C. Tsai, L. Apostol, C. O. Honda, J. Xu, L. M. Wong, T. Zhang, A. Lee, A. Gnanasekar, T. K. Honda, S. Z. Kuo, M. A. Yu, E. Y. Chang, M. “. R. Rajasekaran, W. M. Ongkeko, Using machine learning of clinical data to diagnose COVID-19: a systematic review and meta-analysis, BMC Medical Informatics and Decision Making 20 (1) (sep 2020). doi:10.1186/s12911-020-01266-z.

[5] M. Ambigavathi, D. Sridharan, Analysis of Clustering Algorithms in Machine Learning for Healthcare Data, Springer Singapore, 2020. doi:10.1007/978-981-15-6634-912.

[6] M. R. Aravindakshan, S. K. Maity, A. Paul, P. Chakrabarti, C. Mandal, J. Sarkar, Distinct pathoclinical clusters among patients with uncontrolled type 2 diabetes: results from a prospective study in rural india, BMJ Open Diabetes Research & Care 10 (1) (2022) e002654. doi:10.1136/bmjdrc-2021-002654.

[7] R. N. Bergman, The minimal model of glucose regulation: a biography, in: Mathematical Modeling in Nutrition and the Health Sciences, Springer, 2003, pp. 1–19.

[8] W. Bolie, Coefficients of normal blood glucose regulation, J Appl Physiol. 16(5) (1961) 783–8. doi:10.1152/jappl.1961.16.5.783.

[9] J. Sturis, K. S. Polonsky, E. Mosekilde, E. V. Cauter, Possible mechanisms underlying slow oscillations of human insulin secretion, T. U. of Denmark (1991) 160.

[10] J. Sturis, K. S. Polonsky, E. Mosekilde, E. Van Cauter, Computer model for mechanisms underlying ultradian oscillations of insulin and glucose, American Journal of Physiology - Endocrinology and Metabolism 260 (5 23-5) (1991). doi:10.1152/ajpendo.1991.260.5.e801.

[11] R. Hovorka, V. Canonico, L. J. Chassin, U. Haueter, M. Massi-Benedetti, M. O. Federici, T. R. Pieber, H. C. Schaller, L. Schaupp, T. Vering, M. E. Wilinska, Nonlinear model predictive control of glucose concentration in subjects with type 1 diabetes, Physiological Measurement 25 (4) (2004) 905–920. doi:10.1088/0967-3334/25/4/010.

[12] C. Dalla Man, A. Caumo, C. Cobelli, The oral glucose minimal model: Estimation of insulin sensitivity from a meal test, IEEE Transactions on Biomedical Engineering 49 (5) (2002) 419–429. doi:10.1109/10.995680.

[13] E. D. Lehmann, T. Deutsch, A physiological model of glucose-insulin interaction in type 1 diabetes mellitus, J. Biomed. Eng. 14 (3) (1992) 235–242.

[14] J. D. Elashoff, T. J. Reedy, J. H. Meyer, Analysis of gastric emptying data, Gastroenterology 83 (6) (1982) 1306–1312.

[15] C. Man, M. Camilleri, C. Cobelli, A system model of oral glucose absorption: Validation on gold standard data, IEEE Transactions on Biomedical Engineering 53 (12) (2006) 2472–2478. doi:10.1109/tbme.2006.883792.

[16] A. Misra, Ethnic-specific criteria for classification of body mass index: a perspective for asian indians and american diabetes association position statement, Diabetes technology & therapeutics 17 (9) (2015) 667–671.

[17] M. T. Wentworth, R. C. Smith, H. T. Banks, Parameter selection and verification techniques based on global sensitivity analysis illustrated for an HIV model, SIAM/ASA Journal on Uncertainty Quantification 4 (1) (2016) 266–297. doi:10.1137/15m1008245.

[18] Y. Chu, J. Hahn, Parameter set selection for estimation of non-linear dynamic systems, AIChE Journal 53 (2007) 2858–2870.

[19] E. Walter, L. Pronzato, Qualitative and quantitative experiment design for phenomenological models—a survey, Automatica 26 (2) (1990) 195–213. doi:10.1016/0005-1098(90)90116-y. URL https://doi.org/10.1016/0005-1098(90)90116-y

[20] K. Schittkowski, Parameter estimation in dynamic systems, in: Progress in Optimization, Springer, 2000, pp. 183–204.

[21] J. Guedj, R. Thiébaut, D. Commenges, Maximum likelihood estimation in dynamical models of hiv, Biometrics 63 (4) (2007) 1198–1206.

[22] S. Singer, J. Nelder, Nelder-mead algorithm, Scholarpedia 4 (7) (2009) 2928.

[23] R. Brun, P. Reichert, H. R. Künsch, Practical identifiability analysis of large environmental simulation models, Water Resources Research 37 (4) (2001) 1015–1030.

[24] P. Virtanen, G. et. al., SciPy 1.0 Contributors, SciPy 1.0: Fundamental Algorithms for Scientific Computing in Python, Nature Methods 17 (2020) 261–272. doi:10.1038/s41592-019-0686-2.

[25] R. K. Goyal, V. Cristofaro, M. P. Sullivan, Rapid gastric emptying in diabetes mellitus: Pathophysiology and clinical importance, Journal of Diabetes and its Complications 33 (11) (2019) 107414. doi:10.1016/j.jdiacomp.2019.107414.

[26] L. V. Gromova, S. O. Fetissov, A. A. Gruzdkov, Mechanisms of glucose absorption in the small intestine in health and metabolic diseases and their role in appetite regulation, Nutrients 13 (7) (2021) 2474. doi:10.3390/nu13072474.

[27] K. L. Hoehn, A. B. Salmon, C. Hohnen-Behrens, N. Turner, A. J. Hoy, G. J. Maghzal, R. Stocker, H. V. Remmen, E. W. Kraegen, G. J. Cooney, A. R. Richardson, D. E. James, Insulin resistance is a cellular antioxidant defense mechanism, Proceedings of the National Academy of Sciences 106 (42) (2009) 17787–17792. doi:10.1073/pnas.0902380106.

[28] X. Wang, X. Zhao, R. Zhou, Y. Gu, X. Zhu, Z. Tang, X. Yuan, W. Chen, R. Zhang, C. Qian, S. Cui, Delay in glucose peak time during the oral glucose tolerance test as an indicator of insulin resistance and insulin secretion in type 2 diabetes patients, Journal of Diabetes Investigation 9 (6) (2018) 1288–1295. doi:10.1111/jdi.12834. URL https://doi.org/10.1111/jdi.12834

[29] R. A. Wright, S. Krinsky, C. Fleeman, J. Trujillo, E. Teague, Gastric emptying and obesity, Gastroenterology 84 (4) (1983) 747–751. doi:10.1016/0016-5085(83)90141-5. URL https://doi.org/10.1016/0016-5085(83)90141-5

[30] R. Jumpertz, M. S. Thearle, J. C. Bunt, J. Krakoff, Assessment of non-insulin-mediated glucose uptake: association with body fat and glycemic status, Metabolism 59 (10) (2010) 1396–1401.

